# Cost-effectiveness of implementing HIV and HIV/syphilis dual testing among key populations in Viet Nam: a modeling analysis

**DOI:** 10.1101/2022.02.28.22271651

**Authors:** David Coomes, Dylan Green, Ruanne Barnabas, Monisha Sharma, Magdalena Barr-DiChiara, Muhammad S. Jamil, Rachel Baggaley, Morkor Newman Owiredu, Virginia Macdonald, Van Nguyen, Son Vo Hai, Melanie M. Taylor, Teodora E Wi, Cheryl Johnson, Alison L. Drake

## Abstract

**Objectives:** Key populations, including sex workers, men who have sex with men, and people who inject drugs, have a high risk of HIV and sexually transmitted infections (STIs). We assessed the health and economic impacts of different HIV and syphilis testing strategies among three key populations in Viet Nam using a dual HIV/syphilis rapid diagnostic test (RDT).

**Setting:** We used the Spectrum AIDS Impact Model to simulate the HIV epidemic in key populations in Viet Nam and evaluated five testing scenarios. We used a 15-year time horizon and all costs are from the provider’s perspective.

**Participants:** We include the entire population of Viet Nam in the model.

**Interventions:** We model five testing scenarios among key populations: 1) annual testing with an HIV rapid diagnostic test (RDT), 2) annual testing with a dual RDT, 3) biannual testing using dual RDT and HIV RDT, 4) biannual testing using HIV RDT, and 5) biannual testing using dual RDTs.

**Primary and secondary outcome measures:** The primary outcome is incremental cost-effectiveness ratios (ICERS). Secondary outcomes include HIV and syphilis cases and costs for each proposed intervention.

**Results:** Annual testing using a dual HIV/syphilis RDT was cost saving and averted 3,206 HIV cases and treated 7,719 syphilis cases compared to baseline over 15 years. Biannual testing using one dual test and one HIV RDT, or two dual tests both averted an additional 875 HIV cases and were cost-effective ($1,024 and $2,518 per DALY averted, respectively). Annual or biannual HIV testing using HIV RDTs and separate syphilis tests were more costly and less effective than using one or two dual RDTs.

**Conclusions:** Annual or biannual HIV and syphilis testing using dual RDTs among key populations can be cost-effective and support countries in reaching global reduction goals for HIV and syphilis.

**STRENGTHS AND LIMITATIONS OF THIS STUDY:**

- Strength: Our model presents novel cost-effectiveness estimates for the use of dual HIV/syphilis testing in key populations that can inform health planners
- Strength: We include five testing scale up scenarios using both HIV RDT and dual HIV/syphilis RDT
- Strength: Our model is informed by demographic, behavioral, and biological data from government sources, surveys, surveillance, publicly available reports, databases, and peer-reviewed literature
- Limitation: We made some assumptions regarding the timing and uptake of HIV and syphilis testing among key populations that may be inaccurate.
- Limitation: Our model assumes that increased syphilis testing and treatment will not impact syphilis prevalence, however, it is unknown whether increased testing will reduce or increase syphilis prevalence.

## INTRODUCTION

Key populations, including people who inject drugs (PWID), men who have sex with men (MSM), sex workers (SW), and transgender populations, are at higher risk of acquiring both HIV and syphilis. HIV incidence is significantly higher among key populations in all regions compared to the general population; however, differences vary substantially by region and by key population.[1] While key populations represent 25% of new HIV cases in sub-Saharan Africa, they represent 80% of new HIV cases in the rest of the world.[2] Recent data suggests syphilis incidence, while remaining prevalent in low- and middle-income countries (LMIC), is increasing among key populations, particularly MSM.[1, 3, 4] World Health Organization (WHO) HIV testing guidelines recommend HIV retesting at least annually for key populations and more frequent testing (3-6 months) for those with high ongoing risk.[5] WHO guidelines for syphilis screening depend on population and setting. Rapid diagnostic tests (RDTs) are increasingly being used to screen for syphilis in some settings, yet laboratory-based testing remains common,[6] leaving many key populations unreached by syphilis testing. With the introduction of prequalified dual HIV/syphilis RDTs, and the recent WHO recommendation to offer dual HIV/syphilis testing in antenatal care (ANC) settings,[7] it is important to consider how further integration and expansion of dual HIV/syphilis testing could benefit key populations.

Since 2015, WHO has recommended immediate initiation of antiretroviral therapy (ART) for all people living with HIV (PLHIV) [8] and the United Nations 95-95-95 targets aim to diagnose 95% of PLHIV, provide 95% of PLHIV with ART, and ensure 95% PLHIV on ART are virally suppressed.[9] Despite progress towards these goals – in 2019 81% of PLHIV knew their HIV status and 67% were on ART – this progress is uneven; only 2/3 of key populations are aware of their HIV status.[2] While key populations lag behind the general population in all phases of testing, linkage to treatment, and viral suppression, the largest gap exists in testing.[10] WHO has also developed a global strategy on sexually transmitted infections (STIs) which aims for a 90% reduction in syphilis incidence by 2030, and for 70% of key populations to have access to STI and HIV services, including prevention, testing, and treatment.[11] Increased syphilis testing and treatment may reduce syphilis burden among key and general populations, as well as HIV incidence since early symptomatic syphilis increases risks of HIV acquisition and transmission.[3] Currently, the frequency of syphilis testing recommended by WHO is based on local epidemics; however, the optimal frequency for syphilis testing among key populations is unknown.

In Viet Nam, the national HIV prevalence is <1% in the general population, and significantly higher in key populations, with prevalence ranging between 3-13% among PWID, MSM, and FSW. Similarly, syphilis prevalence among MSM (6.7%) and FSW (2.1%) are also higher than the general population (0.3%).[12] With budgetary constraints in HIV and STI programs, and the health sector, identifying cost-effective strategies for targeted HIV and syphilis testing among high risk groups in Viet Nam is crucial to inform policymakers seeking to optimize resource allocation to maximize population health. We modeled the health impacts and costs associated with varying frequencies of HIV and syphilis testing for key populations, using test scenarios that include a dual HIV/syphilis RDT.

## METHODS

### Settings and Populations

We modeled three key populations: MSM, PWID, and FSW (and their clients) in Viet Nam, using national level HIV prevalence and syphilis prevalence estimates for each key population **(Table 1)**.

**Table 1.**
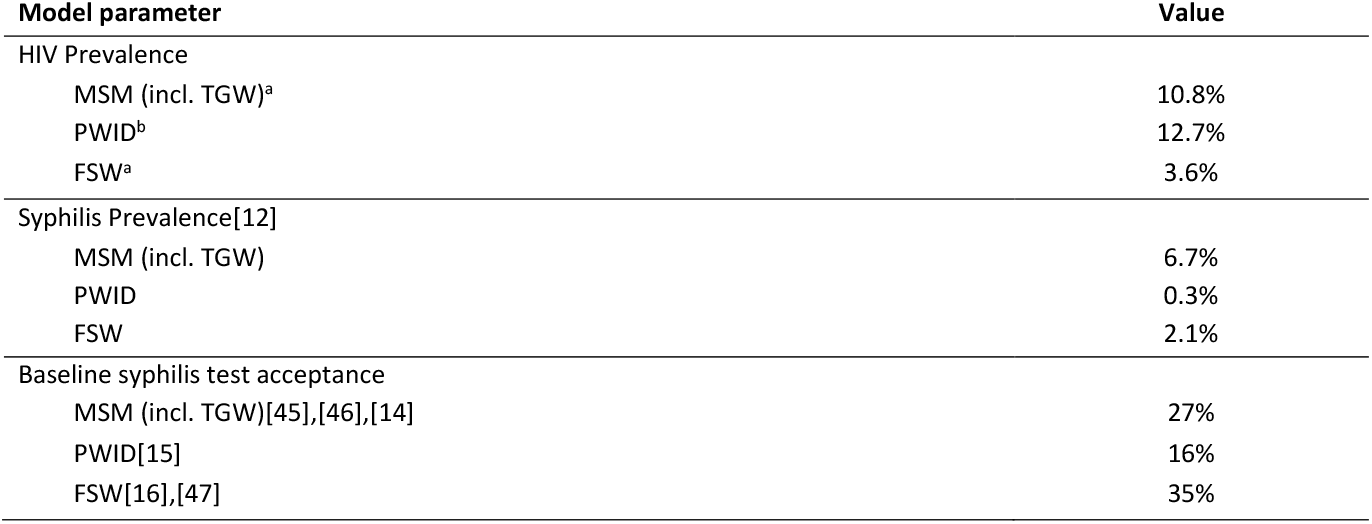
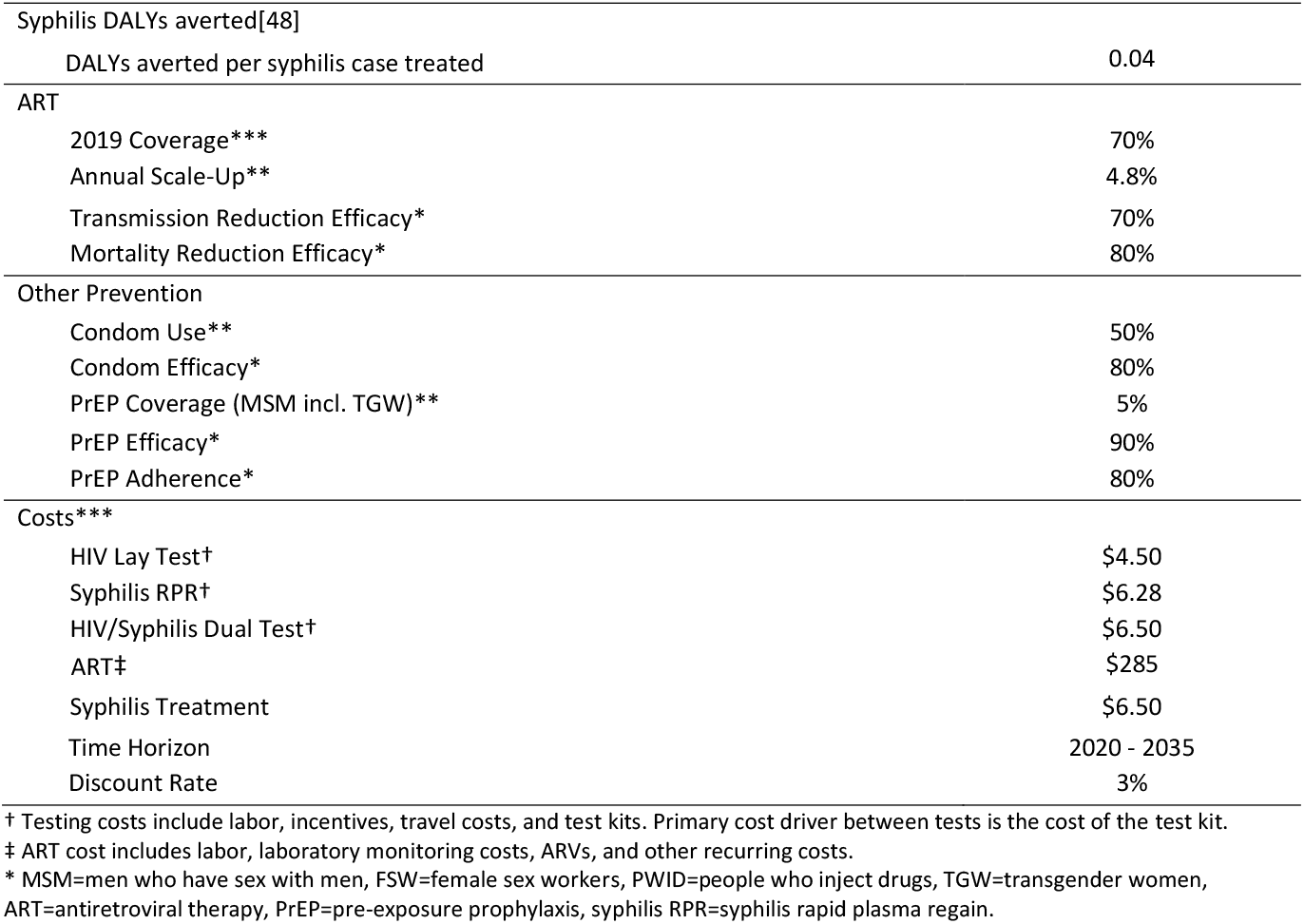
Model parameters for Spectrum input and cost-effectiveness analysis of HIV and syphilis testing scale up among key populations in Viet Nam. ^a^ = 2018 Viet Nam HIV Sentinel Surveillance. ^b^ = 2019 Viet Nam HIV Sentinel Surveillance. * = Spectrum model prior. ** = assumed. *** = Based on information from in-country source.

| Model parameter | Value |
| --- | --- |
| HIV Prevalence |  |
| MSM (incl. TGW)<sup>a</sup> | 10.8% |
| PWID<sup>b</sup> | 12.7% |
| FSW<sup>a</sup> | 3.6% |
| Syphilis Prevalence[12] |  |
| MSM (incl. TGW) | 6.7% |
| PWID | 0.3% |
| FSW | 2.1% |
| Baseline syphilis test acceptance |  |
| MSM (incl. TGW)[45],[46],[14] | 27% |
| PWID[15] | 16% |
| FSW[16],[47] | 35% |
| Syphilis DALYs averted[48] |  |
| DALYs averted per syphilis case treated | 0.04 |
| ART |  |
| 2019 Coverage*** | 70% |
| Annual Scale-Up** | 4.8% |
| Transmission Reduction Efficacy* | 70% |
| Mortality Reduction Efficacy* | 80% |
| Other Prevention |  |
| Condom Use** | 50% |
| Condom Efficacy* | 80% |
| PrEP Coverage (MSM incl. TGW)** | 5% |
| PrEP Efficacy* | 90% |
| PrEP Adherence* | 80% |
| Costs*** |  |
| HIV Lay Test† | \$4.50 |
| Syphilis RPR† | \$6.28 |
| HIV/Syphilis Dual Test† | \$6.50 |
| ART‡ | \$285 |
| Syphilis Treatment | \$6.50 |
| Time Horizon | 2020 - 2035 |
| Discount Rate | 3% |

† Testing costs include labor, incentives, travel costs, and test kits. Primary cost driver between tests is the cost of the test kit.
‡ ART cost includes labor, laboratory monitoring costs, ARVs, and other recurring costs.
\* MSM=men who have sex with men, FSW=female sex workers, PWID=people who inject drugs, TGW=transgender women, ART=antiretroviral therapy, PrEP=pre-exposure prophylaxis, syphilis RPR=syphilis rapid plasma regain.

### Model

We used a deterministic, compartmental model to simulate the HIV epidemic in key populations from 2020-2035 using the AIDS Impact Model within the Spectrum software package (v 5.76). The model estimates annual HIV incidence, AIDS mortality, and disability. We simulated the impact of increasing HIV testing frequency using the Goals model within Spectrum, as previously described.[13] Briefly, the model is age- and sex-stratified with compartments for MSM, PWID, FSW and their clients, and low- and medium-risk heterosexuals.^1^ The model was parameterized with demographic, behavioral, and biological data from government sources, surveys, surveillance, publicly available reports, databases, and peer-reviewed literature. To estimate syphilis burden, we used population size estimates from the Goals model and population-specific estimates of prevalence;[12] we estimate the number of persons tested positive and treated for syphilis infection under each scenario. This model assumes that syphilis testing and treatment does not impact syphilis prevalence. For both HIV and syphilis, disability-adjusted life years (DALYs) are calculated for each scenario. Model key parameters are shown in **Table 1**.

### Costs

Cost inputs include cost per HIV test, initial syphilis test (rapid plasma reagin (RPR)), and the dual HIV/syphilis RDT. We used local data on the personnel, commodities, and transport costs associated with lay testing and estimate costs (**Table 1**). ART costs include personnel, commodities, clinical follow-up, and laboratory monitoring. This analysis includes the costs of intervention delivery and treatment (penicillin and ART) but does not consider additional averted sequelae costs such as the treatment of opportunistic infections due to uncontrolled HIV. All costs are from the provider’s perspective and reported in 2019 US dollars.

### Scenarios

Our baseline scenario estimates annual HIV testing based on current WHO recommendations,[5] and syphilis testing based on observed uptake. In the baseline scenario, we assume 50% of individuals in key populations test annually for HIV, and syphilis testing with RPR occurs at rates specific to each key population (**Table 2**).[14– 16] We considered alternative scenarios with varying testing frequency and test type (separate HIV and syphilis RPR, or a combined dual syphilis/HIV RDT) among key populations from 2020 to 2035. Scenarios modeled include: **1)** annual HIV testing with RDT and baseline syphilis RPR testing, **2)** annual testing with dual RDT, **3)** biannual testing (2 times per year), first with dual RDT and then with HIV RDT, **4)** biannual HIV testing with RDT and baseline syphilis RPR testing, and **5)** biannual testing with dual RDT **(Table 2)**. We assumed 75% test acceptance for the first test in all scenarios except baseline, and 90% of those who accepted the first test would accept the second test in all scenarios that include biannual testing.

**Table 2:**
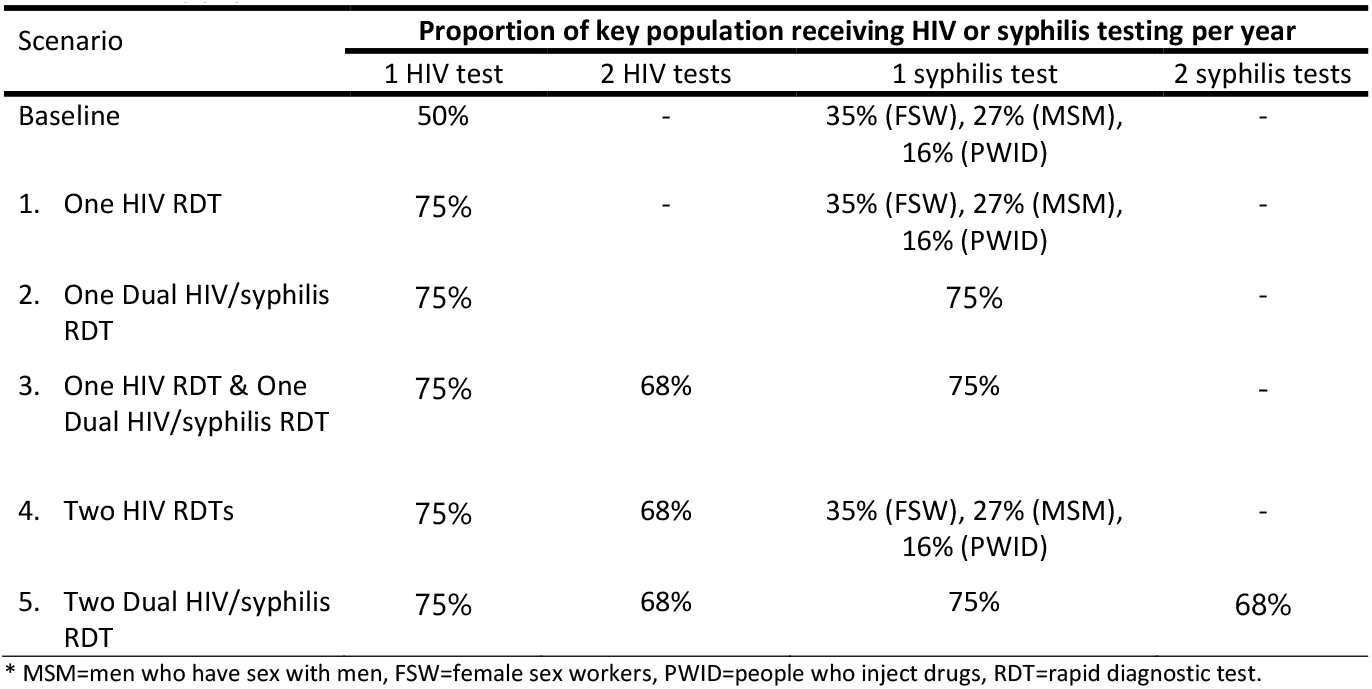
HIV/syphilis testing scenarios among key populations in Viet Nam. The table cells show the proportion of key populations in Viet Nam that receive each test per year. If not specified, the proportion refers to all key populations.

| Scenario | Proportion of key population receiving HIV or syphilis testing per year |  |  |  |
| --- | --- | --- | --- | --- |
|  | 1 HIV test | 2 HIV tests | 1 syphilis test | 2 syphilis tests |
| Baseline | 50% | - | 35% (FSW), 27% (MSM), 16% (PWID) | - |
| 1. One HIV RDT | 75% | - | 35% (FSW), 27% (MSM), 16% (PWID) | - |
| 2. One Dual HIV/syphilis RDT | 75% |  | 75% | - |
| 3. One HIV RDT & One Dual HIV/syphilis RDT | 75% | 68% | 75% | - |
| 4. Two HIV RDTs | 75% | 68% | 35% (FSW), 27% (MSM), 16% (PWID) | - |
| 5. Two Dual HIV/syphilis RDT | 75% | 68% | 75% | 68% |

\* MSM=men who have sex with men, FSW=female sex workers, PWID=people who inject drugs, RDT=rapid diagnostic test.

We modeled increases in testing coverage by adjusting the percent of adults living with HIV (PLWH) on ART. The baseline scenario assumes 95% ART coverage among PLWH by 2028 (4.8% increase per year) based on recent ART scale-up in Viet Nam; test coverage increases by 6.0% per year with annual HIV testing (HIV RDT or dual RDT), and by 7.2% per year with biannual testing. Maximum test coverage is 95% for each model. All models assume ART coverage of 66% of men and 72% of women living with HIV in 2020 based on estimates from the Viet Nam HIV-AIDS Technical Working Group (TWG). We assume universal treatment among those who test positive for syphilis, individuals treated cannot become re-infected within the same year,[17] and no changes to syphilis prevalence under test case scenarios.

### Cost-Effectiveness

Health impact was measured in DALYs averted, HIV infections averted, syphilis infections treated, and AIDS-related deaths averted over the 15-year time horizon. This time horizon was chosen because it reflects current HIV program planning in Viet Nam. Costs and health benefits were discounted at 3% annually per standard health economic evaluations. Incremental costs were calculated as costs incurred and averted by the testing strategy. We utilized WHO guidelines for cost-effectiveness threshold: less than gross domestic product (GDP) per capita is considered cost-effective in Viet Nam ($2,715 USD in 2019).[18]

### Model calibration and sensitivity analyses

Models were calibrated to national HIV prevalence estimates for each key population. Monte Carlo sensitivity analyses were conducted to evaluate robustness of results to changes in: HIV and syphilis testing coverage, scenario program uptake rate, HIV and syphilis testing cost, HIV and syphilis treatment cost, average years on ART, and time horizon. **Table S1** shows the model parameters, ranges, and distributions used in the sensitivity analysis.

### Patient and public involvement

It was not appropriate or possible to involve patients or the public in the design, conduct, reporting, or dissemination plans of our research.

### Ethics approval

This study did not receive nor require ethics approval as it does not involve human or animal participants.

## RESULTS

Increasing annual HIV test coverage from 50% (baseline) to 75% using an HIV RDT (scenario 1) or a dual RDT (scenario 2) is projected to avert 3,206 HIV infections and 660 AIDS-related deaths by 2035 in Viet Nam (**Table 3**). Annual testing using dual RDT led to treatment of an additional 7,719 syphilis cases over 15 years compared to using HIV RDT, but the number of HIV infections averted was the same. HIV testing with either HIV or dual RDT biannually (scenarios 3, 4, & 5) was projected to avert an additional 875 HIV infections and 183 AIDS-related deaths by 2035 compared to annual testing. Testing using a dual HIV/syphilis RDT biannually among key populations is projected to lead to an additional 124,460 syphilis cases treated by 2035, compared to annual testing using a dual RDT (116,680 total syphilis cases treated).

**Table 3.**
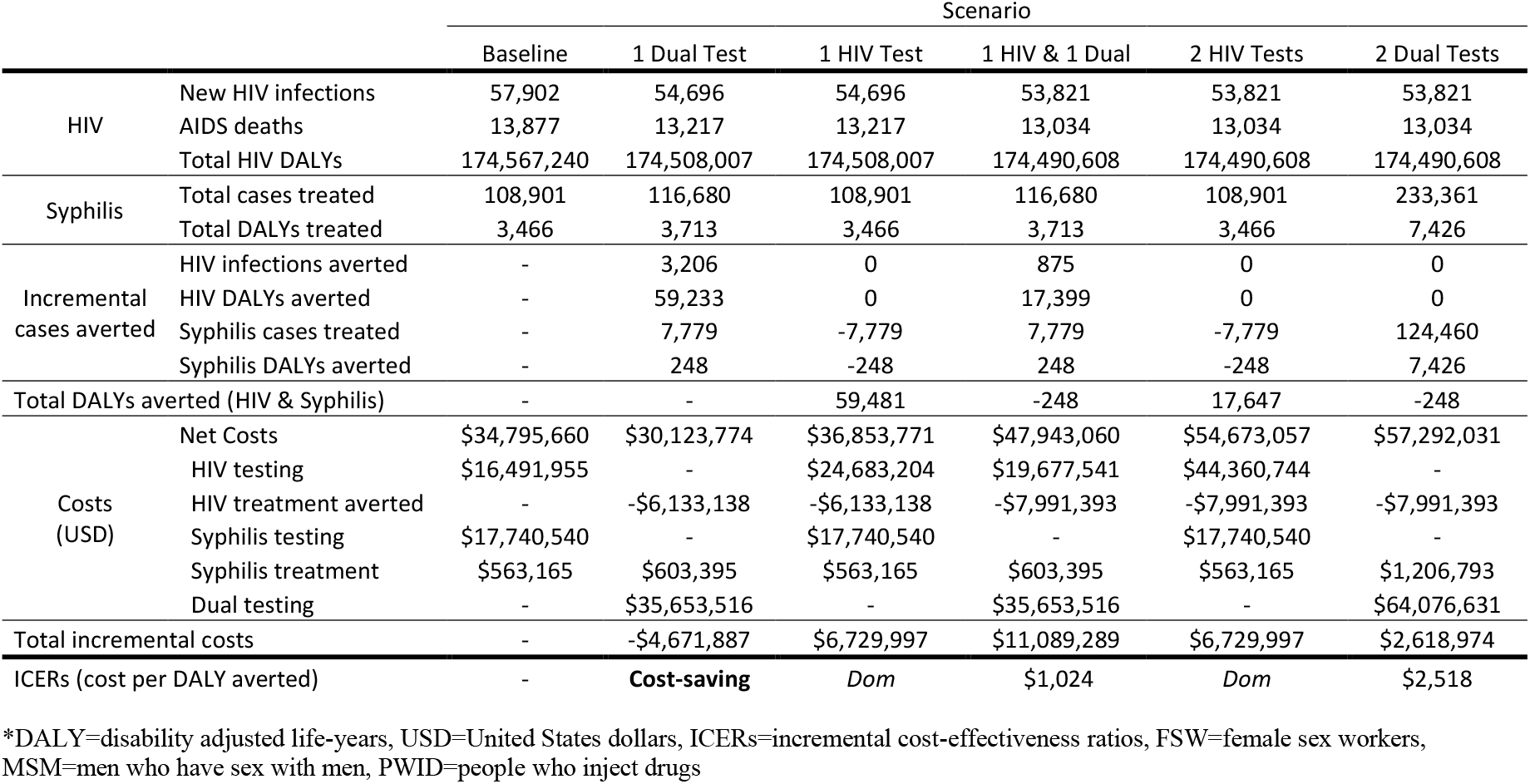
Estimated HIV and syphilis infections, and cost-effectiveness of increased HIV and Dual HIV/syphilis testing among key populations in Viet Nam from 2020-2035. Each scenario refers to the number of tests per year. The baseline scenario assumes that 50% of key populations are tested for HIV each year and syphilis testing rates are specific to each sub-population (FSW, MSM, and PWID). Scenarios including one test per year assume a 75% test acceptance rate, and those that include two tests per year assume a 75% test acceptance rate for the first test, and a 68.5% test acceptance rate for the second test. *Incremental cases averted, Total DALYs averted*, and *ICERs* compare each scenario to the previous one.

|  |  | Scenario |  |  |  |  |  |
| --- | --- | --- | --- | --- | --- | --- | --- |
|  |  | Baseline | 1 Dual Test | 1 HIV Test | 1 HIV & 1 Dual | 2 HIV Tests | 2 Dual Tests |
| HIV | New HIV infections | 57,902 | 54,696 | 54,696 | 53,821 | 53,821 | 53,821 |
|  | AIDS deaths | 13,877 | 13,217 | 13,217 | 13,034 | 13,034 | 13,034 |
|  | Total HIV DALYs | 174,567,240 | 174,508,007 | 174,508,007 | 174,490,608 | 174,490,608 | 174,490,608 |
| Syphilis | Total cases treated | 108,901 | 116,680 | 108,901 | 116,680 | 108,901 | 233,361 |
|  | Total DALYs treated | 3,466 | 3,713 | 3,466 | 3,713 | 3,466 | 7,426 |
| Incremental cases averted | HIV infections averted | - | 3,206 | 0 | 875 | 0 | 0 |
|  | HIV DALYs averted | - | 59,233 | 0 | 17,399 | 0 | 0 |
|  | Syphilis cases treated | - | 7,779 | -7,779 | 7,779 | -7,779 | 124,460 |
|  | Syphilis DALYs averted | - | 248 | -248 | 248 | -248 | 7,426 |
| Total DALYs averted (HIV & Syphilis) |  | - | - | 59,481 | -248 | 17,647 | -248 |
| Costs (USD) | Net Costs | \$34,795,660 | \$30,123,774 | \$36,853,771 | \$47,943,060 | \$54,673,057 | \$57,292,031 |
| | HIV testing | \$16,491,955 | - | \$24,683,204 | \$19,677,541 | \$44,360,744 | - |
| | HIV treatment averted | - | -\$6,133,138 | -\$6,133,138 | -\$7,991,393 | -\$7,991,393 | -\$7,991,393 |
| | Syphilis testing | \$17,740,540 | - | \$17,740,540 | - | \$17,740,540 | - |
| | Syphilis treatment | \$563,165 | \$603,395 | \$563,165 | \$603,395 | \$563,165 | \$1,206,793 |
| | Dual testing | - | \$35,653,516 | - | \$35,653,516 | - | \$64,076,631 |
| Total incremental costs | | - | -\$4,671,887 | \$6,729,997 | \$11,089,289 | \$6,729,997 | \$2,618,974 |
| ICERs (cost per DALY averted) | | - | **Cost-saving** | *Dom* | \$1,024 | *Dom* | \$2,518 |

\*DALY=disability adjusted life-years, USD=United States dollars, ICERs=incremental cost-effectiveness ratios, FSW=female sex workers, MSM=men who have sex with men, PWID=people who inject drugs

The most effective strategy was biannual testing with the dual RDT, which was projected to avert 4,081 HIV infections (7% of total infections), 76,632 HIV DALYs (0.04% of total HIV DALYs), and treat 124,460 cases of syphilis by 2035 compared to the baseline scenario. The discounted cost of implementing this scenario over 30 years is $57.3 million USD compared to $34.8 million USD for the baseline scenario. The testing cost of implementing biannual testing using the dual RDT is almost four times the cost of baseline testing with an HIV RDT ($64.1 million vs. $16.5 million, respectively), but an estimated $8.0 million USD in HIV treatment costs would be saved by biannual HIV testing, and $17.7 million USD in syphilis testing costs would be averted by using the dual RDT. The cost of biannual testing with an HIV RDT and continuing to test for syphilis with RPR is slightly higher than biannual testing with the dual RDT ($57.3 vs. $54.7 million USD, respectively), but the latter strategy treats an estimated 124,000 more cases of syphilis over 15 years.

Annual testing with the dual RDT is cost saving compared to the baseline scenario (**Figure 1**). Annual HIV testing with RDT is more expensive and averts fewer DALYs than with the dual RDT (strongly dominated). The next most efficient scenario is biannual testing using one dual RDT and one HIV RDT, which is cost-effective ($1,024 USD per DALY averted). Biannual testing with the dual RDT is also cost-effective ($2,518 USD per DALY averted) and more cost-effective than using the HIV RDT (weakly dominated). Despite slightly higher initial costs, the discounted cost of annual testing with a dual RDT becomes less than that of current testing within two years, due to decreased ART costs associated with HIV averted (**Figure S1**).

**Figure 1.**
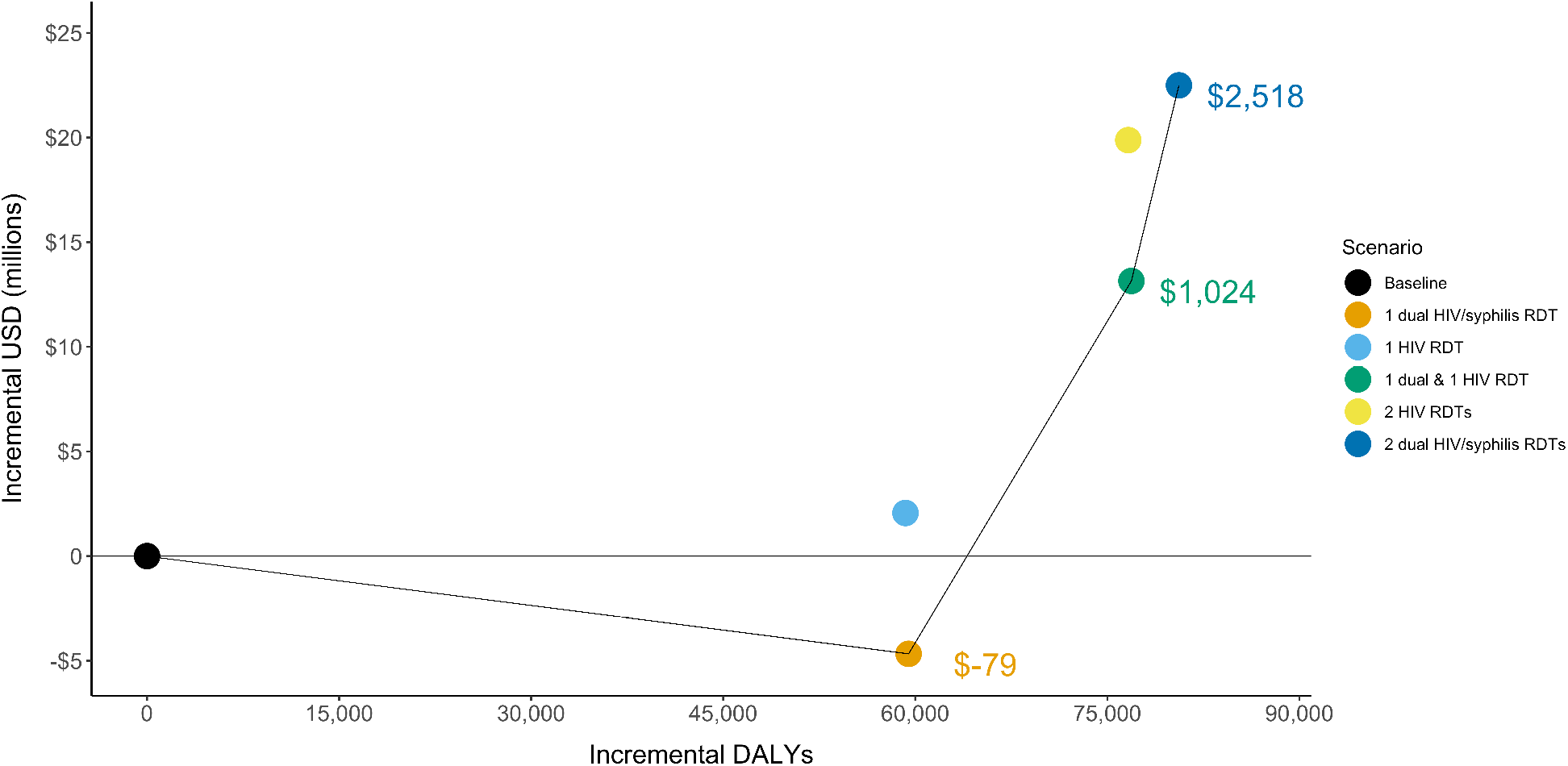
Efficiency frontier presenting the total disability adjusted life years (DALYs) and costs for five testing scenarios among key populations. The solid line indicates the scenarios that are not dominated by other scenarios. Dominated indicates that a scenario is either more costly and less effective or has a higher ICER than a scenario that is more effective. The ICERs for the non-dominated scenarios are shown. DALYs=disability adjusted life-years, ICER=incremental cost-effectiveness ratio, RDT=rapid diagnostic test, USD=United States dollars

Sensitivity analyses including all scenarios found that an annual dual RDT (scenario 2) is cost-saving in 59% of the simulations and either cost-saving or cost-effective (at $2,715 per DALY averted) in all simulations (**Table S2**). In non-dominated scenarios (scenarios 2, 3, and 5), using dual RDT annually or biannually was cost-effective or cost-saving in most simulations, while biannual testing with one dual RDT and one HIV RDT was cost-saving in only 31% of simulations (**Figure 2**). In univariate sensitivity analysis adjusting costs, our scenarios that involve one dual RDT (scenarios 2 and 3) remain cost-effective even after all costs (testing and treatment) are increased by 50%. In this sensitivity analysis, biannual dual testing is no longer cost-effective ($3,777 per DALY averted).

**Figure 2.**
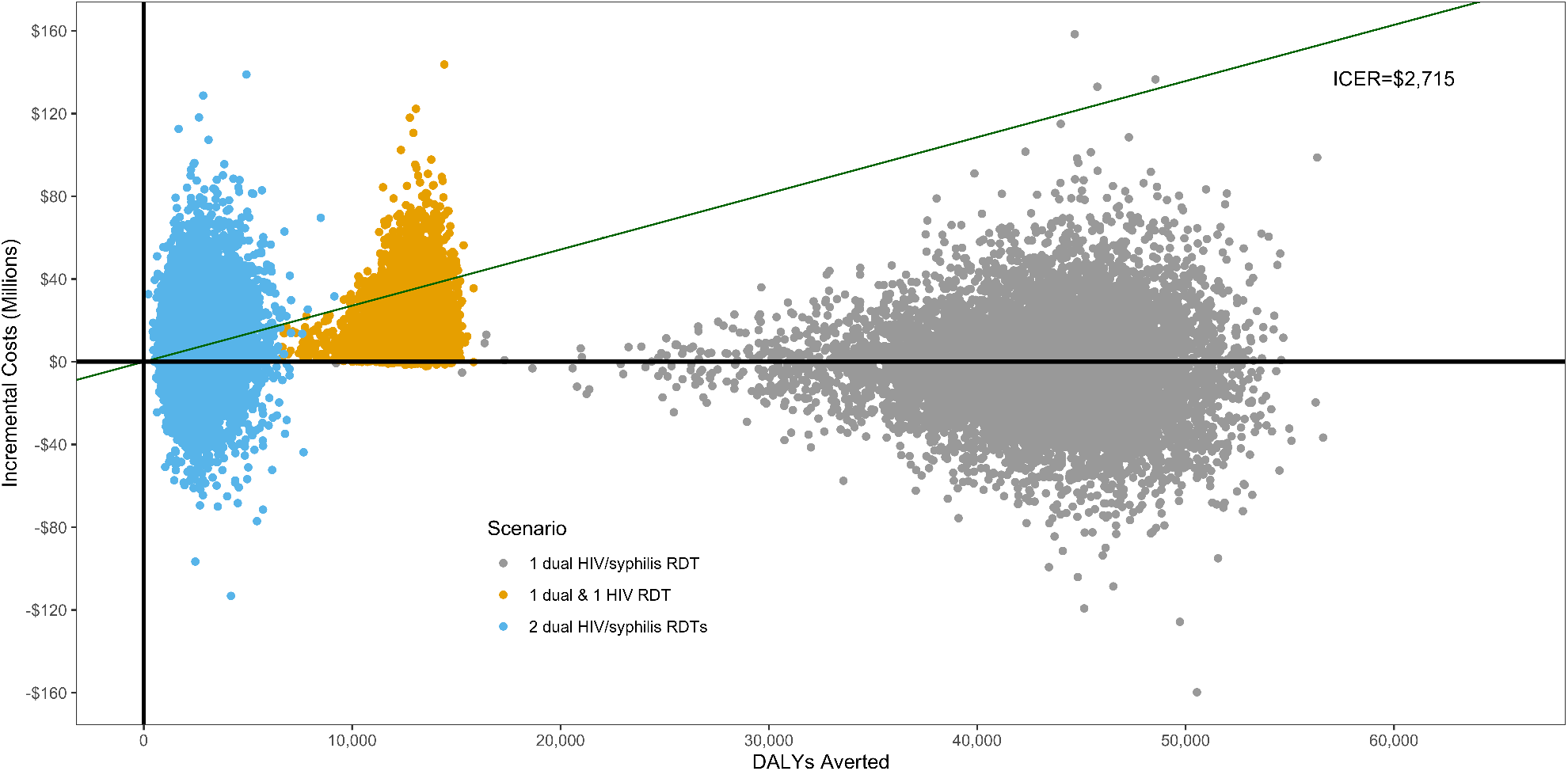
Sensitivity analysis of non-dominated scenarios using a Monte Carlo simulation of the cost effectiveness of HIV/syphilis dual testing among key populations in Viet Nam. Plot shows 10,000 iterations in which 17 key parameters were randomly adjusted. All points below the green line are cost-effective at $2,715 per DALY averted and those below the solid black line (y-intercept) are cost-saving. Only non-dominated scenarios are shown in this figure; cost-effectiveness of *1 Dual Test* is compared to baseline, *1 HIV Test & 1 Dual Test* is compared to *1 Dual Test*, and *2 Dual Tests* is compared to *1 HIV Test & 1 Dual Test*. DALYs=disability adjusted life-years, ICER=incremental cost-effectiveness ratio, RDT=rapid diagnostic test, USD=United States dollars.

## DISCUSSION

In our model, we found that implementing annual testing with the dual RDT at 75% coverage was cost-saving, averted more HIV infections, and treated more syphilis cases compared to annual testing using HIV RDT at 50% coverage and current syphilis testing in Viet Nam. While biannual testing with one dual RDT and one HIV RDT was projected to be more costly, it would avert more HIV and syphilis related DALYs and using dual RDT for both tests would avert additional DALYs attributed to syphilis. Increasing the frequency of HIV testing to one or two tests per year using only HIV RDTs, while continuing to test for syphilis using RPR, was not efficient compared to other strategies.

Implementing biannual testing substantially increases testing costs, but also prevents more HIV infections, therefore averting more ART-related costs. Increasing test frequency may be cost-saving or cost-effective although it incurs considerable costs in the near term while costs averted may not be observed for many years. Annual testing using a dual RDT can help offset some near-term costs as it is less expensive than using HIV RDT and syphilis RPR. Policymakers must weigh the health impact and cost-effectiveness of different testing scenarios over time against current affordability given HIV and syphilis testing budgets; however, using the dual RDT will help integrate syphilis testing within existing HIV testing programs, improving program efficiencies.[19]

Implementation of dual RDT is occurring in some settings; preliminary reports indicate that 48% of countries use dual RDT in ANC, and 25% use dual RDT in key populations, although the extent of this use is unknown (WHO HIV Testing Services, 2021). PEPFAR and the Global Fund cover dual RDT in ANC,[20] and there are multiple dual RDTs qualified by the Global Fund and WHO.[21] The use of dual RDT during ANC could be a model for improving HIV/STI integration among those at high risk for both HIV and syphilis, such as key populations, however, there are multiple operational challenges associated with using dual tests, namely that of integrating HIV and STI programs.[22]

Benefits of the dual test are its cost-effectiveness and potential to reach more at-risk individuals. Annual or biannual testing can enable earlier identification of HIV-positive individuals for faster ART initiation and prevention of onward transmission. Annual HIV testing for key populations is recommended by WHO, and more frequent testing (every 3-6 months) may be advised for those with individual risk factors, including those using pre-exposure prophylaxis (PrEP) and key populations presenting with STIs.[23] Individuals presenting with syphilis symptoms should also test for HIV, and using the dual RDT is less costly as compared to a syphilis RPR and HIV RDT. As policy makers scale up PrEP among key populations in Viet Nam, including at least one dual RDT in the testing algorithm may be more cost-effective than using HIV RDTs alone. In addition, using dual RDT tests can facilitate lay providers to offer both HIV and syphilis testing for their community.[24]

Our results were robust to sensitivity analyses, suggesting that testing annually or biannually using dual RDTs remains cost-effective if testing costs increase and HIV prevalence decreases. In scenarios involving dual RDT, the majority (>98%) of benefits, as measured in DALYs, come from averting HIV infections rather than treating syphilis due to the relatively larger burden of disease from HIV than syphilis. However, since the cost of a dual RDT is only slightly higher than the cost of an HIV RDT, it is cheaper to use a dual RDT than separate HIV RDT and syphilis RPR tests.

Increased HIV testing can reduce HIV-associated morbidity and mortality and transmission from PLHIV through early detection and initiation of ART. While models suggest high ART coverage would result in substantial declines in HIV incidence,[25, 26] empiric data from countries with population-level viral suppression exceeding 73% (e.g. Australia, eSwatini, and Thailand) have observed less significant reductions in HIV incidence relative to predictions from mathematical models.[27] Similarly, when high ART coverage was achieved in a series of cluster-randomized trials in sub-Saharan Africa, it resulted in decreased population-level HIV incidence; however, this decrease was insufficient to end HIV as a public health threat.[28–31] These discrepancies may be attributed to delayed diagnosis and ART initiation following infection,[32, 33] and gaps in the 95-95-95 targets for some population groups, for example young men and key populations. More frequent HIV testing strategies could increase earlier diagnosis and initiation on ART, and focusing testing and linkage efforts on key populations could reduce the access and coverage disparities in these groups. However, more frequent testing will also increase program costs, not only through additional commodity procurement, but also for health systems, program coordination, and outreach. In settings of concentrated HIV epidemics, health planners may benefit from targeting limited testing resources towards high-risk groups such as key populations.

Dual RDTs may also increase syphilis testing frequency and coverage among key populations who are more likely to access HIV testing. Previous research has shown that coupling rapid syphilis testing in ANC may also increase HIV test coverage in LMICs, particularly in settings where HIV test coverage is low.[34] This strategy may be similarly effective at increasing test coverage for both diseases among key populations, as well as augment current ANC testing by reaching women in key populations who present late or are missed by ANC services. While there is a lack of data on dual RDTs among key populations, models of dual RDT during ANC have been shown to be cost-saving or cost-effective among both key populations and the general population of pregnant women.[7, 35] While dual RDTs are likely more effective in the context of ANC since testing can avert more adverse outcomes associated with congenital syphilis and mother-to-child HIV transmission, we find dual RDTs may also be cost-effective among non-pregnant key populations.

Our results are consistent with previously published models that show expanded testing and early access to ART for key populations in Viet Nam will cost-effectively reduce the country’s HIV burden.[36, 37] Additionally, models from both low- and high-resource countries suggest HIV testing every 3-6 months among key populations can be cost-effective in concentrated epidemics.[38][39] However, HIV risk within key populations is not homogenous; further targeting of higher-risk groups within key populations may be needed to achieve efficient testing regimens. While we examine the impact of increased testing frequency among key populations as a whole, previous research has described the benefits of targeting high-risk groups within key populations.[40] Individuals who engage in risky behaviors, such as those with more sexual partners, practicing unprotected sex, or needle sharing may benefit from additional testing or linkage to HIV prevention such as PrEP. Further research is needed on the optimal testing intervals for higher-risk groups of key populations.

Approximately one-third of key populations are not aware of their HIV status. Programs focusing on HIV testing and treatment among FSW and PWID in Viet Nam have shown success in reducing HIV prevalence; however, less than a third of MSM reported testing for HIV in 2015, likely contributing to increases in HIV prevalence among MSM in the past decade.[41] Annual syphilis testing among key populations in Viet Nam is similarly low, ranging from 16% among PWID to 36% among FSW. [14–16] Due to high dual prevalence of HIV and syphilis among key populations, dual testing is a promising strategy to increase testing coverage and linkage to care.

Our analysis has several limitations. We did not include the cost of scaling-up and training providers in administering dual RDTs. However, RDT are easy to use and can be administered by a lay provider, and rapid results can minimize loss to follow-up. Overall, dual RDTs have been shown to have adequate performance in field settings in Viet Nam among key populations.[42] Dual RDTs may also increase HIV test coverage as they can be easily conducted by community health workers outside of healthcare settings, and they may be more acceptable to some members of key populations who are concerned about stigma associated with testing.[43] Despite this, some additional training, supervision, and support will be needed to scale-up dual RDT use among key populations.

Some model assumptions regarding the timing of HIV and syphilis testing may be inaccurate. We assumed in scenarios that included a dual RDT, additional syphilis RPR tests would not be conducted. PLWH who know their status and present for syphilis screening do not need an HIV test. We assume regular testing intervals for the entire population in each scenario, but it is possible people who engage in risky behaviors or experience symptoms may seek more frequent retesting than biannually. We did not include the increased costs of outreach to achieve increased test coverage of key populations. Considerable expansions of first time testing among MSM in Viet Nam have recently been achieved through social media campaigns, perhaps providing a guide for cost-effectively increasing testing uptake among key populations.[41, 44] We also did not consider the burden that increasing the testing coverage and frequency may have on the health system; however, as testing may be conducted effectively using lay providers, increased testing may not substantially impact the provision of other services.[43] Although targeting key populations in lower prevalence regions may be more difficult and costly, these results are robust to increased costs and it will likely remain an effective use of resources. We assume that syphilis screening will not impact syphilis prevalence rates. Increased screening may reduce prevalence by increasing early treatment, but syphilis screening also has the potential to increase prevalence as individuals with latent syphilis are unlikely to transmit the infection to others unless they are treated and then infected again. Finally, there is limited data on population size, HIV and syphilis prevalence, and health seeking behaviors among key populations. We based our model input on estimates included in published literature as well as Viet Nam country sources.

Since data on the impact of retesting on population HIV incidence is limited, we made conservative assumptions about the frequency of linkage to care and ART use following retesting. We assumed that HIV testing frequency would increase in Viet Nam among key populations in the baseline scenario but testing frequency would increase more quickly under the other scenarios. Because of this, we believe our estimates of the impact of increased testing frequency are conservative.

Our study suggests that annual or biannual HIV and syphilis testing among key populations in Viet Nam using a dual RDT will increase HIV and syphilis detection and treatment, while remaining cost-saving or cost-effective. Integrating HIV and other STI testing can streamline services as well as expand testing and help countries with epidemics concentrated in key populations reach 95-95-95 targets.

## Data Availability

All data produced in the present study are available upon reasonable request to the authors

## CONTRIBUTORSHIP STATEMENT

CJ and AD devised the project and the main conceptual ideas. DC, DG, and RB parameterized the model. DC and DG carried out the model implementation. VH and SVH provided model feedback. All authors provided critical feedback and helped shape the research, analysis, and manuscript.

## DATA SHARING STATEMENT

Extra data is available by emailing David Coomes

## Footnotes

1 Low-risk heterosexuals are those in stable couples while medium-risk heterosexuals are those that engage in casual sex but are not in a high-risk group (high risk groups: MSM, FSW, PWID).

