## Supplemental figures for "Cost-effectiveness of implementing HIV and HIV/syphilis dual testing among key populations in Viet Nam: a modeling analysis"

**Table S1. Parameters and probability distributions for Monte Carlo simulation.** Table shows the baseline model parameter values and the probability distributions used for random draws of 17 variables for 10,000 Monte Carlo simulations. Beta distributions were used for all proportion parameters. For the beta distribution, the alpha and beta parameters were calculated as the baseline value multiplied by 100, except for the *impact* parameter where an alpha and beta of 25 was used. Gamma distributions were used for all other parameters. For the gamma distribution, the alpha parameter was calculated as the square of the baseline parameter divided by the square of the standard deviation. The beta parameter was calculated as the square of the standard deviation divided by the baseline parameter.

| Model Parameter | Baseline value | Distribution | St. Dev | alpha/beta |
|---|---|---|---|---|
| **Baseline HIV test acceptance** | 50% | Beta | N/A | 50 |
| **Recommended test acceptance** | 75% | Beta | N/A | 75 |
| **Additional test drop off rate** | 10% | Beta | N/A | 10 |
| **HIV Lay Test Cost** | $4.50 | Gamma | $3.00 | N/A |
| **Syphilis RPR1 Cost** | $6.28 | Gamma | $3.50 | N/A |
| **HIV/Syphilis Dual Test Cost** | $6.50 | Gamma | $3.50 | N/A |
| **ART Treatment Cost** | $285 | Gamma | $50 | N/A |
| **Syphilis Treatment Cost** | $6.50 | Gamma | $3.50 | N/A |
| **Avg Years on ART** | 25 | Gamma | 3 | N/A |
| **Time Horizon** | 2035 | Gamma | 3 | N/A |
| **Impact** | 100% | Beta | N/A | 25 |
| **Baseline Syphilis test acceptance FSW** | 35% | Beta | N/A | 35 |
| **Baseline syphilis test acceptance MSM** | 27% | Beta | N/A | 27 |
| **Baseline syphilis test acceptance PWID** | 16% | Beta | N/A | 16 |
| **Baseline syphilis prevalence FSW** | 2.1% | Beta | N/A | 2.1 |
| **Baseline syphilis prevalence MSM** | 6.7% | Beta | N/A | 6.7 |
| **Baseline syphilis prevalence PWID** | 0.3% | Beta | N/A | 0.3 |

**Table S2. Sensitivity analysis of all scenarios using a Monte Carlo simulation.** Table shows the percentage of simulations (10,000 iterations) in which each scenario is cost-effective (at $500 or $2,715 per DALY averted), cost-saving, or less-effective. Less effective scenarios are both less effective and more costly as compared to the scenario above. Scenarios are arranged in order of increasing cost and each scenario is compared to the one immediately above; 1 Dual HIV/syphilis RDT is compared to the baseline scenario.

| Scenario | Cost-effective ($500) | Cost-effective ($2,715) | Cost-saving | Less effective |
|---|---|---|---|---|
| 1 Dual HIV/syphilis RDT | 87% | 100% | 59% | 0% |
| 1 HIV RDT | 11% | 12% | 11% | 69% |
| 1 Dual HIV/syphilis RDT & 1 HIV RDT | 45% | 87% | 31% | 0% |
| 2 HIV RDTs | 11% | 12% | 11% | 69% |
| 2 Dual HIV/syphilis RDTs | 49% | 55% | 48% | 0% |

DALY=disability adjusted life-year, RDT=rapid diagnostic test

**Figure S1. Cost pressure analysis of testing scenarios.** Figure shows the discounted cost over time of each scenario. Costs are discounted 3% with a time horizon from 2020 – 2035. Baseline costs include testing costs assuming that 50% of key populations are tested for HIV each year and syphilis testing rates are specific to each sub-population (FSW, MSM, and PWID), and syphilis treatment costs. All other scenarios include the cost of HIV treatment averted compared to the baseline scenario, testing costs, and syphilis treatment costs. Scenarios including one test per year assume a 75% test acceptance rate, and those that include two tests per year assume a 75% test acceptance rate for the first test, and a 68.5% test acceptance rate for the second test. Each scenario refers to the number of tests per year. RDT=rapid diagnostic test, USD=United States dollars

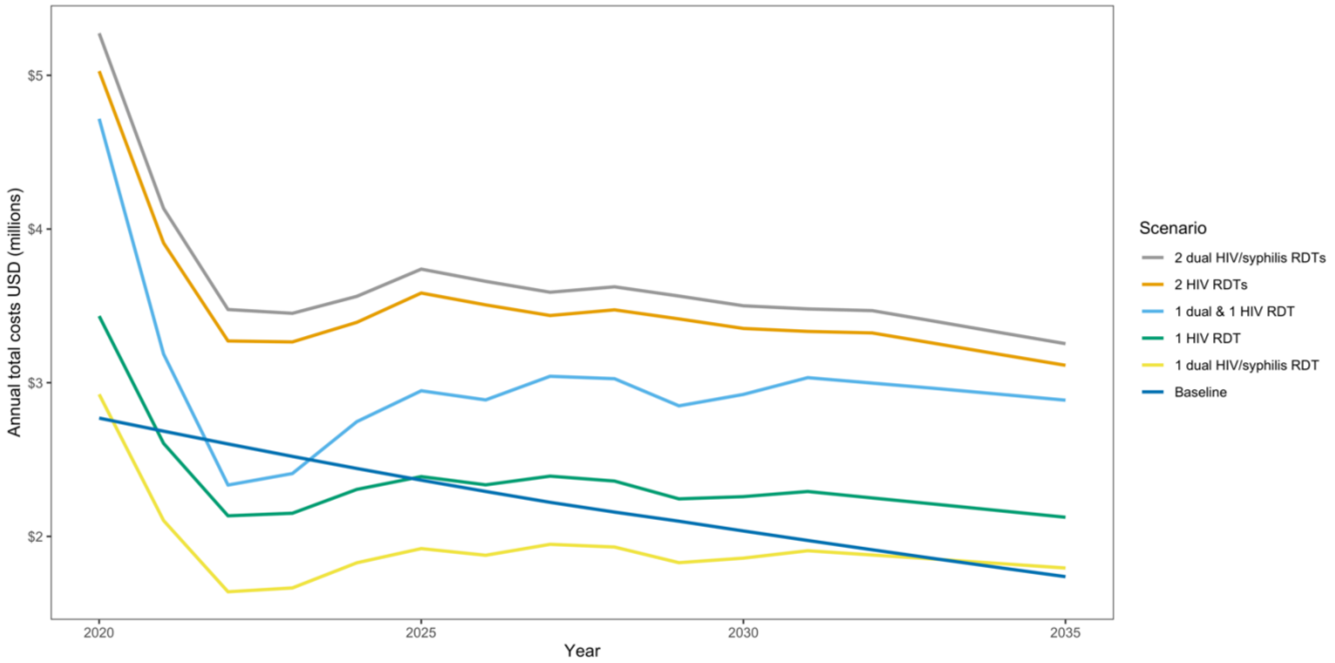
